# Clinical Features of Hemodialysis (HD) patients confirmed with Coronavirus Disease 2019 (COVID-19): a Retrospective Case-Control Study

**DOI:** 10.1101/2020.07.06.20147827

**Authors:** Huan Zhou, Xiaofen Xiao, Xiaohui Wang, Xianhua Tan, Xin Zhang, Yong He, Jing Li, Guosheng Yang, Mingmei Li, Duan Liu, Shanshan Han, Haibo Kuang

**Author notes:** Correspondence to Xiaohui Wang and Xianhua Tan., Xiaohui Wang, Department of Nephrology, Wuhan No.5 Hospital, Wuhan, Hubei Province, 430050, China, Xianhua Tan, Department of Radiology, Wuhan No.5 Hospital, Wuhan, Hubei Province, 430050, China. Contributed equally (Huan Zhou, Xiaofen Xiao).

## Abstract

**Background:** Since December 2019, Coronavirus Disease 2019(COVID-19) occurred in wuhan, China, and outbreaked rapidly into a global pandemic. This current poses great challenges to hemodialysis (HD) patients.

**Objective:** To make a comprehensive evaluation and comparison between HD patients confirmed with COVID-19 and the general HD patients.

**Methods:** HD patients confirmed with COVID-19 in Wuhan No.5 Hospital were admitted as confirmed group from Jan 10 to Mar 15, 2020. And HD patients not infected in our dialysis center were chosen as control group. General characteristics, laboratory indicators were retrospectively collected, analyzed and compared.

**Results:** A total of 142 cases were admitted, including 43 cases in confirmed group and 99 in control group. Body mass index (BMI) was slightly lower in confirmed group than that in control group (*P*=0.011). The proportion of one or less underlying disease in confirmed group(51.16%) was higher than that in control group(14.14%)(*P*< 0.001), and the proportion of three or more underlying diseases in confirmed group(11.63%) was lower than that in control group(52.53%)(*P*< 0.001). Patients in confirmed group exhibited significantly lower hemoglobin, lymphocyte count, and lymphocyte percentage, but higher neutrophil percentage, neutrophil-to-lymphocyte ratio (NLR), C-reactive protein, aspartate transaminase, and alkaline phosphatase. There was no significant difference in age, gender, dialysis age, primary disease, the using of ACEI/ARB, platelet-to-lymphocyte ratio (PLR), and other indicators between the two groups.

**Conclusions:** Faced with Severe Acute Respiratory Syndrome-CoV-2 (SARS-CoV-2), HD patients with lower BMI and hemoglobin were more susceptible to be infected, which might be related to malnutrition. Once confirmed with COVID-19, HD patients expressed obviously dis-regulated of inflammation and immune.

## Introduction

Since December 2019, a novel coronavirus outbreak occurred in wuhan, China[1, 2, 3]. On February 12, 2020, the coronavirus panel of the international committee on the classification of viruses recommended that novel coronavirus be officially named “SARS-CoV-2”. On the same day, the World Health Organization (WHO) announced that the disease was officially named Coronavirus Disease 2019 (COVID-19)[4]. As an acute respiratory infectious disease, COVID-19 has been incorporated into the law of the People’s Republic of China on the prevention and treatment of infectious diseases. On March 11, 2020, WHO officially declared the novel coronavirus into a global pandemic. Wuhan, as one of the most seriously affected areas, is now out of the woods. But the global number of confirmed and death cases is raising. By the end of April 24, 2020,the cumulative number of infected cases was more than 2.8 million cases, and 197,003 deaths globally.

Previous reports about the clinical characteristics of diagnosed patients in China alerted that the severity of COVID-19 became more serious in elder people and those with underlying medical condition(s) [5, 6]. This current poses great challenges to HD patients. On average, HD patients are elder and have a large numbers of comorbidities. Most of them must travel to and from the hospital for hemodialysis two or three times a week. And simultaneously, patients in the dialysis centers are relatively dense and mobile. So it is urgently necessary to pay more attention to this group of people. so far, epidemiological data in patients on HD patients confirmed COVID-19 during SARS-CoV-2 outbreak is limited, except for some single cases reported.

In our study, a comprehensive evaluation and comparison between HD patients confirmed with COVID-19 and the general HD patients were implemented. We aimed to have a deeper understanding about the clinical features of HD patients after infected with SARS-CoV-2.

## Methods

### Study design and patients

This was a single-centre, retrospective, observational case-control study. HD patients diagnosed with COVID-19 from Jan 10 to Mar 15, 2020, were admitted as confirmed group. We followed the clinical diagnosis criteria for COVID-19 in the New Coronavirus pneumonia Prevention and Control Program by the National Health Commission of China [1,7, 8]. At the same time, the ordinary HD patients who were not infected in our dialysis center at the hospital were chosen as control group. All of them received regular dialysis (3 times per week, 4 hours each time) in our dialysis center. Patients owning complete clinical data were selected in our research, and eliminated those infected of HBV, HCV, HIV, syphilis, and other pathogens. Finally the number of cases were determined: 43 cases in confirmed group and 99 cases in control group.

### Data collection

Data of confirmed group was collected by consulting electronic medical records and asking patients themselves or their family members. Data of control group was obtained by logging into Chinese National Renal Data System (CNRD). The basic information included: age, gender, height, weight, dialysis age, primary disease, underlying disease, the using of angiotensin converting enzyme inhibitor/ angiotensin receptor blockers (ACEI/ ARB) in the past one month. Laboratory indicators included white blood cell count, hemoglobin, neutrophil count, neutrophil percentage, lymphocyte count, lymphocyte percentage, platelet count, neutrophil-to-lymphocyte ratio (NLR), platelet-to-lymphocyte ratio (PLR), C-reactive protein, alanine transaminase, aspartate transaminase, total bilirubin, total protein, albumin, and alkaline phosphatase. BMI is calculated by height and weight: weight (kg) / height^2^ (m^2^). In our study, we divided the patients into 3 categories according to the kinds of underlying disease: ≤1 (one or less kind), and 2 (two kinds), ≥3(three or more kinds).

### Statistical analysis

Statistical analyses were done using the SPSS software (version 25.0). All statistical tests were conducted by bilateral test, with 95% confidence interval. We present categorical variables as count (%), and continuous variables as mean ± SD if they were normally distributed or median (IQR) if they were not. We compared means for continuous variables by using independent group *t* tests when the data were normally distributed; otherwise, we used the Mann-Whitney U test. Categorical variables were compared by the *χ2* test or Fisher’s exact test. A two-sided of less than 0.05 was considered statistically significant.

## Results

### General characteristics of confirmed HD patients and control patients

A total of 142 cases were admitted. The general information of the two groups was shown in the Table 1, including 43 cases in confirmed group and 99 cases in control group. There were significant differences in BMI and underlying disease between the two groups. In the confirmed group, BMI was slightly lower than that in control group (P=0.011). The proportion of one or less underlying disease in confirmed group(51.16%) was higher than in control group(14.14%)(χ 2=21.71, P< 0.001), and the proportion of three or more underlying disease in confirmed group(11.63%) was lower than that in control group(52.53%)(χ 2=20.86, P< 0.001). While the proportion of two underlying diseases between them was no difference (P= 0.655).

**Table 1.** General characteristics of the confirmed HD patients and the control patients

| Characteristics | Confirmed cases<br>(n=43) | Control cases<br>(n=99) | Total<br>(n=142) | *T/Z/χ<sup>2</sup>*<br>value | *P* value |
| --- | --- | --- | --- | --- | --- |
| Age (years) | 60.81 ± 10.65 | 60.75 ± 11.83 | 60.77 ± 11.45 | 0.032 | 0.975 |
| Sex: |  |  |  | 0.071 | 0.789 |
| Male | 29(67.44) | 69 (69.70) | 98 (69.01) |  |  |
| Female | 14(32.56) | 30 (30.30) | 44 (30.99) |  |  |
| BMI(kg/m<sup>2</sup>) | 20.28(19.03-21.83) | 21.40(20.07-23.93) | 21.09(19.36-23.38) | -2.546 | 0.011 |
| Dialysis age (mo) | 32.00(8.00-60.00) | 34.00(16.00-82.00) | 33.00(15.00-70.50) | -1.787 | 0.074 |
| Primary disease: |  |  |  | 3.698 | 0.296 |
| Glomerulonephritis | 20(46.51) | 36(36.36) | 56(39.44) | 1.293 | 0.256 |
| Hypertensive<br>nephropathy | 10(23.26) | 33(33.33) | 43(30.28) | 1.442 | 0.23 |
| Diabetic nephropathy | 13(30.23) | 26(26.26) | 39(27.46) | 0.237 | 0.626 |
| Others* | 0(0.00) | 4(4.04) | 4(2.82) | 1.788 | 0.315 |
| Kinds of underlying<br>disease: |  |  |  | 28.371 | <0.001 |
| ≤1 | 22(51.16) | 14(14.14) | 36(25.35) | 21.71 | <0.001 |
| 2 | 16(37.21) | 33(33.33) | 49(34.51) | 0.199 | 0.655 |
| ≥3 | 5(11.63) | 52(52.53) | 57(40.14) | 20.86 | <0.001 |
| ACEI/ARB |  |  |  | 0.059 | 0.807 |
| Yes | 17(39.53) | 37(37.37) | 54(38.03) |  |  |
| No | 26(60.47) | 62(62.63) | 88(61.97) |  |  |

other\* obstructive nephropathy(n=2) uric acid nephropathy(n=1) the polycystic kidney(n=1).
Data are mean ± SD, n (%), or median (IQR). *P* values comparing groups are from Mann-Whitney U test, and *t* test for categorical variables, ANOVA for continuous variables.
Abbreviations: BMI, Body mass index; ACEI/ARB, Angiotensin-Converting Enzyme Inhibitors/ Angiotensin Receptor Blocker.

The average age was 60.77 ± 11.45 year and the median dialysis age was 33.00 (15.00-70.50) months. Male was more predominant in two groups, in which 29(67.44%) males confirmed and 69(69.7%) males control (P=0.975). Glomerulonephritis (56; 39.44%) was the most common primary disease, followed by hypertensive nephropathy (43; 30.28%), diabetic nephropathy (39; 27.46%), and others (as obstructive nephropathy, uric acid nephropathy, the polycystic kidney) (4; 2.82%). 17(39.53%) patients in confirmed group were taking the ACEI/ARB, and 37(37.37%) cases in control group, which expressed no difference. There was no statistically significant difference between the two groups in terms of age, gender, dialysis age, primary disease and the using of ACEI/ARB.

### Laboratory characteristics of confirmed group and control group

We observed substantial differences in laboratory findings between confirmed group and control group (Table 2). Patients in confirmed group exhibited significantly lower hemoglobin, lymphocyte count, and lymphocyte percentage, and higher neutrophil percentage, NLR, C-reactive protein, aspartate transaminase, and alkaline phosphatase. Other indicators were not significantly different between the two groups.

**Table 2.** Laboratory characteristics of the confirmed HD patients and the control

| Laboratory characteristics | Confirmed group (n=43) | Control group (n=99) | Total (n=142) | T/Z/$\chi^2$ value | P value |
| --- | --- | --- | --- | --- | --- |
| White blood cell count (*10<sup>9</sup>/L) | 6.14(4.24-7.05) | 5.56(4.20-7.03) | 5.70(4.23,7.04) | -0.815 | 0.415 |
| Hemoglobin (g/L) | 93.40±21.78 | 106.15±20.18 | 102.29±21.42 | -3.379 | 0.001 |
| Neutrophil count (*10<sup>9</sup>/L) | 4.84(3.20-5.33) | 3.99(3.11-5.18) | 4.18(3.13-5.30) | -1.503 | 0.133 |
| Neutrophil percentage (%) | 76.00(72.90-84.10) | 72.2(68.30-76.50) | 73.80(68.78-78.60) | -3.215 | 0.001 |
| Lymphocyte count (*10<sup>9</sup>/L) | 0.70(0.40-1.12) | 0.94(0.77-1.26) | 0.90(0.65-1.24) | -2.824 | 0.005 |
| Lymphocyte percentage (%) | 14.10(10.10-17.60) | 17.80(14.90-22.80) | 16.55(13.27-21.53) | -3.681 | <0.001 |
| Platelet count(*10<sup>9</sup>/L) | 155.35±72.65 | 170.06±66.38 | 165.61±68.42 | -1.179 | 0.24 |
| NLR | 5.36(3.87-8.38) | 4.08(3.03-5.15) | 4.42(3.16-5.97) | -3.277 | 0.01 |
| PLR | 11.01(7.70-16.15) | 9.18(6.39-13.46) | 9.83(6.74-14.22) | -1.716 | 0.086 |
| C-reactive protein (mg/L) | 25.20(7.70-71.00) | 3.50(1.64-7.80) | 5.30(1.95-20.08) | -5.692 | <0.001 |
| Alanine transaminase (U/L) | 10.00(7.00-22.00) | 8.00(7.00-12.00) | 9.00(7.00-13.00) | -1.641 | 0.101 |
| Aspartate transaminase (U/L) | 17.00(13.00-26.00) | 14.00(12.00-16.00) | 14.00(12.00-18.00) | -3.462 | 0.001 |
| Total bilirubin (umol/L) | 4.50(2.90-7.13) | 3.84(2.50-5.55) | 4.08(2.60-5.75) | -1.372 | 0.17 |
| Total protein (g/L) | 69.87±7.89 | 70.89±5.71 | 70.58±6.43 | -0.771 | 0.44 |
| Albumin (g/L) | 36.63±6.27 | 38.52±3.13 | 37.95±4.39 | -1.884 | 0.065 |
| Alkaline phosphatase (U/L) | 60.00(50.00-72.00) | 53.00(40.00-71.00) | 55.00(41.00-71.25) | -2.014 | 0.044 |

Data are mean ± SD, n (%), or median (IQR). P values comparing groups are from $\chi^2$ , Fisher's exact test, or Mann-Whitney U test, t test for categorical variables, and ANOVA for continuous variables.
Abbreviations: NLR, neutrophil-to-lymphocyte ratio; PLR, platelet-to-lymphocyte ratio.

## Discussion

This is the first retrospective case-control study to comprehensively compare clinical features between HD patients confirmed COVID-19 and those general HD patients. In our study, there were indeed many differences in two groups, by the observation and comparison between 43 cases confirmed and 99 cases controlled. These differences were distributed in general data and laboratory index.

As we all known, the understanding of COVID-19 is gradually improved, and the diagnosis and treatment program is deepened from the 1st to the 7th. Due to low virus titers, sampling at late stage of illness, and inappropriate swabbing sites, false negative cases might be common for COVID-19 infection cases at special period[9]. In order to reduce rate of misdiagnosis, clinical diagnosis was introduced in the 5th guidelines[7]. Of 43 confirmed patients in our study, 19(44.19%) were with positive RT-PCR Tests for SARS-CoV-2, and 24 (55.81%) were diagnosed with COVID-19 through clinical diagnosis. These clinical diagnosis cases were mainly judged by epidemiological history, clinical manifestations and chest CT scan results, which could ensure their accuracy of diagnosis.

BMI is closely related to patients with Chronic Kidney Disease (CKD). A prospective cohort study showed that maintaining a normal BMI can slow the development of CKD, while abnormal BMI can accelerate the progression of CKD to End-stage Renal Disease (ESRD) [10]. Similarly, in HD patients, together with hemoglobin and serum albumin, BMI usually represents the nutritional status of HD patients, which is one of the main factors affecting the prevalence of various diseases and the survival rate in HD patients. In our study, BMI of patients in confirmed group was significantly lower than that of control group *(P*=0.011). Meanwhile, in confirmed group, hemoglobin and albumin were lower than that in control group. Therefor, we believe that HD patients with lower BMI and hemoglobin were more susceptible to be infected with SARS-CoV-2, which might be related to malnutrition.

The differences in laboratory index between confirmed and control groups were obviously, including lymphocyte count, lymphocyte percentage, neutrophil percentage, NLR, C-reactive protein, aspartate transaminase, and alkaline phosphatase. Lymphocytopenia is a main feature of critical patients with coronavirus infection, such as SARS-CoV and MERS-CoV [11-13]. Elevated inflammatory biomarkers, like C-reactive protein, interleukin-1, interleukin-6 and ferritin, usually due to the hyperinflammatory response or secondary infection. Similar to the acute phase of SARS-CoV infection in humans, SARS-CoV-2 infection also have a hyperinflammatory response and “cytokine storm” in acute phase, which leads to acute respiratory distress syndrome, multiple organ failure, and was the leading cause of death [14-17]. This phenomenon also exists in HD patients. In our study, contrasted with general HD patients, lymphocytes count (*P* = 0.005) and lymphocytes percentage (*P* < 0.001) were lower, and CRP (*P* < 0.001) higher in those confirmed after admission. Previous studies have confirmed that, once patients infected with SARS-CoV-2, NLR was significantly increased and could be used as one of the intuitive indicators for clinical evaluation of the severity of COVID-19 [18]. In our study, compared with control group, NLR was significantly increased in confirmed group(*P* = 0.01). Research suggests that the higher PLR of patients during treatment had longer average hospitalization days [19]. However, there was no significant difference of PLR in HD patients, which may be related to thrombocytopenia in HD patients[20, 21].

At the same time, neutrophil percentage existed higher in confirmed group in our study. This phenomenon may be caused by secondary bacterial infections[22]. It has been recognized that patients of COVID-19 has been damaged with other organs such as liver, heart and kidney[23]. In this study, we also found that AST and ALP were significantly higher than the control group, which may be related to other organ damage after SARS-CoV-2 infection.

Similar to MERS-CoV, the risk of infection SARS-CoV-2 in elders and people with underlying conditions such as chronic obstructive pulmonary disease (COPD), diabetes, hypertension, and chronic cardiovascular disease (CVD) is increased [24-27]. In our study, there existed no significant in age difference between confirmed group and control group, so as to moderate and severe cases in confirmed group (data was not yet available). This may be related to the age composition ratio of dialysis patients themselves. In our study, more underlying disease does not mean higher morbidity. We speculate that because of more attention from their family, organizations, and themselves, HD patients with more underlying diseases decrease their frequency of exposure to SARS-CoV-2, but that is not the case for HD patients with less underlying diseases. While the percentage of male was higher in the two groups, there was no difference in sex ratio between the two groups (*χ*^2^=0.071, *P*=0.789). It has been previously reported that the risk of other diseases, such as pneumonia and tumor, increased with the extension of dialysis age in HD patients[28], which is not obviously in the incidence of COVID-19 (*P*=0.074). As we known, SARS-CoV-2 is related to angiotensin-converting enzyme II (ACE2), which is widely distributed in multiple organs such as lung, heart, kidney and gastrointestinal tract [29,30]. Upregulation of the ACE2-RAAS pathway is well-known to counter-regulate the proinflammatory and profibrotic effects of the ACE-Ang II-AT1 receptor axis in experimental models of HTN and CVD [31-34]. Therefore, it has been speculated that HD patients who have been or are currently using ACEI/ARB are more likely to be infected with SARS-CoV-2 and have a worse prognosis. However, in our study, ACEI/ARB did not significantly increase the incidence and mortality of COVID 19 in HD patient(*χ*^2^=0.059, *P*=0.857). Therefore, age, sex, comorbidity, dialysis age and the using of ACEI/ARB cannot be used as key indicators in assessing the COVID-19 susceptibility factors of HD patients in our study.

Last but not least, in order to avoid the interference of clinical symptoms and laboratory indicators, HD patients infected with other pathogen were specifically excluded. In fact, as a special population, the majority of them have a variety of complications, such as microinflammatory, heart failure, pulmonary edema, and lung infection[28, 35], resulting in complicated clinical manifestations and variable chest CT findings. This requires the medical staff in the dialysis center to master the basic data of all the patients in their own center. At the same time, under the premise of perfect protection, it is one of the necessary protective measures to take the screening of nucleic acid testing and chest CT scans for every hemodialysis patient in the dialysis center.

Our study is limited. Because this is a retrospective case-control study from one single center, and has a small sample, which could be subject to recall bias and selection bias. In conclusion, faced with SARS-CoV-2, HD patients with lower BMI and hemoglobin were more susceptible to be infected, which might be related to malnutrition. Once confirmed with COVID-19, HD patients expressed obviously dis-regulated of inflammation and immune. At the same time, we found that age, gender, comorbidities, dialysis age, and the using of ACEI/ARB could not be used as indicators of susceptibility to COVID-19 in HD patients. Expecting that our study could be helpful to regions currently suffering from the ravages of COVID-19.

## Data Availability

All data generated or analyzed during this study are included in this article.

## Acknowledgments

We thank all patients and their families involved in the study.

## Statement of Ethics

This study was approved by the Medical Ethics Committee of Wuhan No.5 Hospital (Approval Number YJ2020002) and complied with the *Declaration of Helsinki*. Written informed consent was waived due to the rapid emergence of this infectious disease.

## Author Contributions

Huan Zhou, Xiaofen Xiao, Xiaohui Wang and Xianhua Tan conceived and designed the study. Jing Li, Guosheng Yang, Mingmei Li, Duan Liu, Shanshan Han, Haibo Kuang collected the epidemiological and clinical data. Huan Zhou, Xiaofen Xiao, Xin Zhang and Yong He summarised all data. Huan Zhou drafted the manuscript. Xiaohui Wang revised the final manuscript.

## Data Availability Statement

All data generated or analyzed during this study are included in this article.

